# T-cell responses to acute cardiorespiratory or resistance exercise in cohorts of physically active or physically inactive older adults: A randomized complete crossover

**DOI:** 10.1101/2021.04.25.21255567

**Authors:** Rachel M. Graff, Kristofer Jennings, Emily C. LaVoy, Victoria E. Warren, Brad W. Macdonald, Yoonjung Park, Melissa M. Markofski

## Abstract

Aging is associated with many chronic diseases that are maintained and perpetuated by immune dysregulation and chronic systemic inflammation. T-cells often undergo age-related changes, including an accumulation of memory cells, which places individuals at increased risk for novel infections and may predispose them to increased inflammation. Regular exercise training has been suggested to offset age-related changes in T-cells, but the majority of literature is derived from cardiorespiratory exercise (CRE) studies. Much less is understood about the T-cell response to resistance exercise (RE). The purpose of this study was to examine the effects of acute CRE and acute RE on the T-cell response among a cohort of physically active older adults (PA) compared to a cohort of physically inactive older adults (PI).

**METHODS:** Twenty-four healthy older adults (PA n=12; PI n=12; mean ± SD; age (yrs) PA 62 ± 5, PI 64 ± 5; height (cm) PA 170.9 ± 6.9, PI 162.9 ± 8.0; weight (kg) PA 69.3 ± 10.2, PI 68.2 ± 12.8; BMI (kg/m^2^) PA 23.9 ± 3.0, PI 25.6 ± 3.5) completed one bout of CRE and one bout of RE in a randomized order, both at a moderate intensity, and separated by at least 7 days. Blood samples drawn pre-exercise, post-exercise, and 1h post-exercise (recovery) were analyzed for CD4+ and CD8+ T-cells and their differentiation status using surface markers CD45RA, CD62L, and CD57, as well as for Th17 cells (CD4+ CD161+ CD196+) using flow cytometry.

**RESULTS:** PI had higher numbers of circulating CD57+ EMRA CD4+ T-cells (PA, mean ± SE, 1 ± 2 cells/uL; PI, 6 ± 2 cells/uL; p=0.01; z=2.32) than PA at pre-exercise. Both CRE and RE elicited a significant mobilization of highly-differentiated (CD45RA+ CD62L-; CD57+ CD45RA+ CD62L-) CD8+ T-cells into the circulation post-exercise in both PA and PI groups. Furthermore, CRE resulted in a decrease in the number of circulating Th17 cells post-exercise, while RE increased Th17 cell mobilization compared to the CRE response.

**CONCLUSION:** Taken together, T-cells in PA and PI respond similarly to acute CRE and support previously reported data showing a significant mobilization of highly differentiated T-cells. The present study confirms that moderate intensity RE also elicits this response, but highlights potential differences between CRE and RE on the immune responses of T-cells, particularly in PI individuals.

**Clinical trial registration:** This research study was registered at clinicaltrials.gov NCT03794050

## Introduction

Aging is associated with a host of chronic diseases, such as cardiovascular disease, hypertension, cancer, arthritis, type 2 diabetes, and osteoporosis [1]. Age-associated immune system dysregulation (i.e. immunosenescence) and chronic systemic inflammation serve as common underlying conditions that contribute to the development and the perpetuation of many of these diseases. Further, chronic systemic inflammation and these diseases are often exacerbated by physical inactivity, which also increases with aging [2-4].

Within the lymphocyte population, T-cells serve as one of the lymphocyte subtypes that undergo an age-related change. For example, aging has been linked to a reduction in circulating naïve T-cells, which is often accompanied by an accumulation of memory T-cells with higher differentiation status [5]. This shift in the T-cell repertoire reduces the diversity of the T-cell pool and places individuals at greater risk for impaired immunity [6]. Furthermore, highly-differentiated T-cells preferentially secrete pro-inflammatory cytokines such as TNF and IFN-γ, which may also contribute to chronic inflammation and poorer immunity [5].

Physical exercise can alter various lymphocyte subpopulations, particularly those with phenotypes associated with higher effector functions, migration potential, and inflammatory properties such as NK cells, CD8+ T-cells, and gamma-delta T-cells [7, 8]. Within the CD8+ T-cell population, those with a highly-differentiated phenotype are mobilized into the circulation at preferentially higher proportions than their lower-differentiated counterparts, due largely to increases in hemodynamic shear stress and catecholamine secretion that accompany acute exercise [9]. For example, CD8+ T-cells can be classified by differentiation status according to their expression of surface receptors CD45RA and CD62L as naïve (CD45RA+CD62L+), central memory (CM; CD45RA-CD62L+), effector memory (EM; CD45RA-CD62L-), and effector memory RA (EMRA; CD45RA+CD62L-). In response to acute exercise, the proportions of EMRA CD8+ T-cells exhibit the largest increase, followed by EM, CM, and naïve CD8+ T-cells, respectively, without any significant exercise induced changes among naïve, CM, EM, or EMRA CD4+ T-cells [9].

The effects of chronic exercise on lymphocyte and lymphocyte subpopulations are less well understood. Cross-sectional reports indicate that fitness status is a better predictor of senescent T-cell proportions than age [10], which is supported by a longitudinal study that demonstrated that 12 weeks of exercise training increased the number of T-cells that express the cell surface marker CD28 (a marker of low-differentiation status) [11]. Nevertheless, more research is needed in order to determine whether engaging in regular physical activity can reduce or prevent the accumulation of highly-differentiated T-cells and allow for the production or maintenance of the naïve and low-differentiated T-cell pools.

Furthermore, the majority of the literature to date on the effects of exercise on T-cells and T-cell subpopulations is derived from cardiorespiratory exercise studies, with little known about the effects of both acute and chronic resistance training [12]. It is believed that resistance training exerts similar effects to cardiorespiratory training on immune parameters [12], but to our knowledge, cardiorespiratory and resistance conditions have not been directly compared in a crossover study design.

Results from longitudinal studies support the idea that although chronic exercise training may not exert substantial effects on the number of circulating leukocytes rest, [11] in response to acute exercise at any given intensity the absolute number of cells mobilized into the circulation is often lower among trained individuals compared to untrained individuals. The smaller absolute response of leukocytes in trained individuals may be a result of differences in β2-adrenergic receptor sensitivity, glucocorticoid receptor sensitivity, or cell adhesion molecules, each of which may be affected by regular exercise [13].

Therefore, the purpose of the present study was to examine the effects of acute cardiorespiratory and acute resistance exercise on the T-cell response among a cohort of physically active older adults (PA) compared to a cohort of physically inactive older adults (PI).

We hypothesized that both cardiorespiratory and resistance exercise bouts would elicit a T-cell mobilization along with a preferential mobilization of highly-differentiated CD8+ T-cells, but that the magnitude of this mobilization would be higher following cardiorespiratory exercise.

Furthermore, we hypothesized that the magnitude of T-cell and T-cell subpopulation mobilization would be higher among physically inactive older adults compared to physically active older adults.

## Methods

### Research participants

Older adults (55-75 years old) were recruited for this research study. Prior to the first in-person visit, the potential participants were pre-screened to be either PA or PI (Figure 1.1). Physical activity classification was defined using the American College of Sports Medicine guidelines [14]. In order to be considered PA, participants had to be engaging in planned, purposeful exercise training, as defined by participating in at least 20 minutes of vigorous intensity cardiorespiratory activity 3 days per week or 30 minutes of moderate intensity cardiorespiratory activity five days per week, or some combination of vigorous and moderate for at least 3 days per week for the past 3 months. In addition, PA participants engaged in resistance training for all of the major muscle groups approximately 2 days per week for the past 3 months. Participants classified as PI were exercise untrained, which was defined as participating in less than 30 minutes of moderate to vigorous intensity cardiorespiratory physical activity no more than twice per week and no regular resistance training for at least the last 3 months [15]. All participants provided a signed written statement of informed consent prior to enrollment in the study and all study procedures were approved by the Institutional Review Board at the University of Houston. Recruitment and enrollment began January 2019 until target number of participants completed in October 2019.

**Figure 1:**
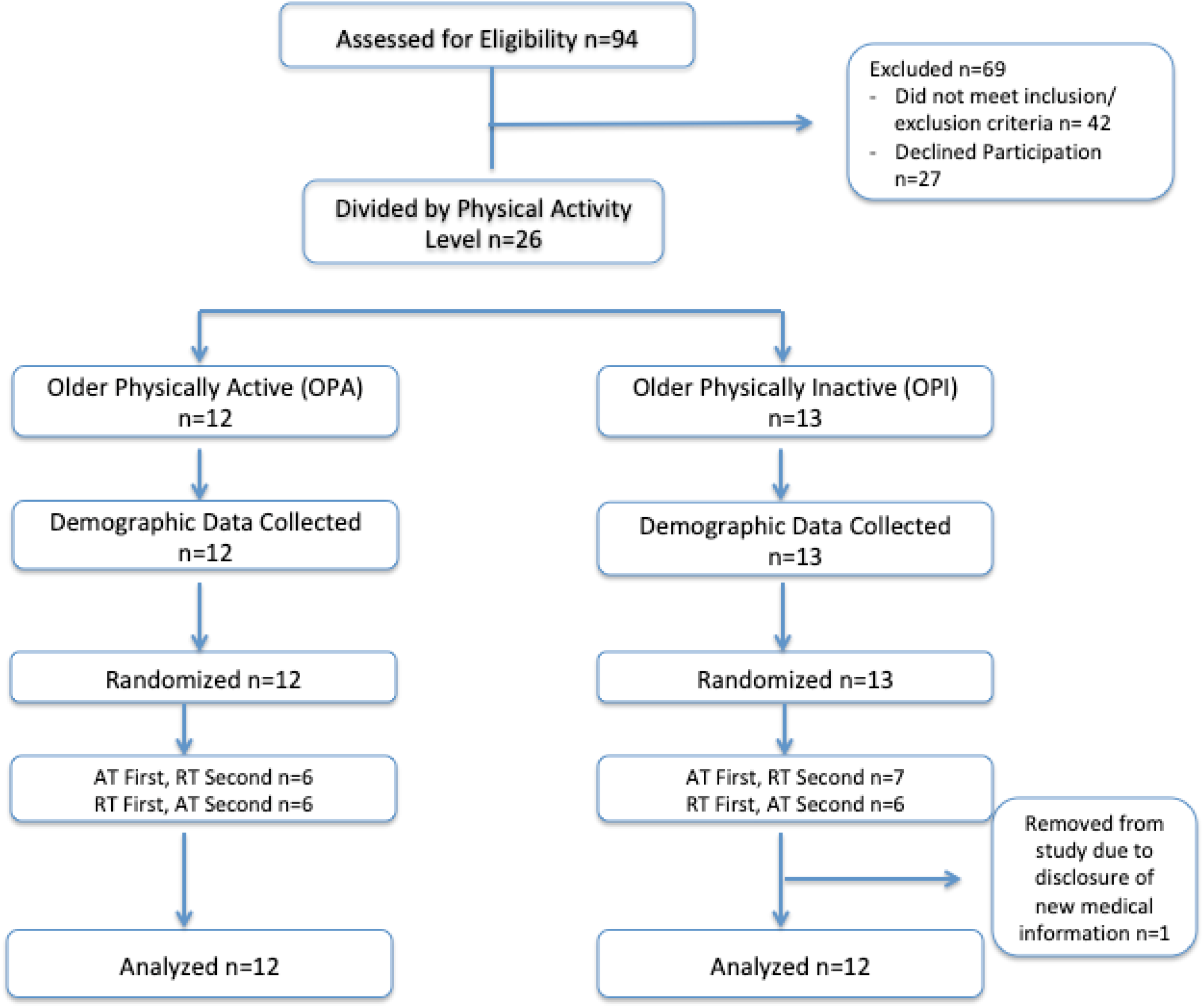
Recruitment and Retention Flow Chart. Twenty four participants completed the study

Participants were excluded if they had: any contraindications to moderate-vigorous exercise; any recent illness or have been instructed not to exercise by their healthcare provider; range of motion restrictions that would prevent them from participating in cardiorespiratory or resistance exercises with proper form (they must be ambulatory); were taking medications (prescription or over the counter) known to influence immune function (including daily NSAID’s and beta blockers), cholesterol-lowering medications (statins), drugs that increase bone mass (bisphosphonates), or steroids; known cardiovascular, respiratory, metabolic, or renal disease, with the exception of controlled hypertension (as defined by resting BP below 140/90) and/or controlled asthma (self-reported); if they fell outside of a BMI range of (18.5 – 30 kg·m^-2^); consumed alcohol or recreational drugs within 24h prior to visits; scheduling conflicts that would prevent them from completing the study. In addition, the women were not pre-menopausal (defined as having a menses in the previous 12 months).

Participants were assigned to PA or PI groups based on the results of the Exercise Frequency Questionnaire, where participants were required to detail the types of weekly activity that they participate in and the length of time that each activity is performed. To confirm activity status, participants were given an ActiGraph accelerometer (wGT3X-BT, ActiGraph ® Corp, Pensacola, FL, USA) along with a set of written instructions, and asked to wear it for seven full days. Participant characteristics are presented in Table 1.1.

**Table 1:**
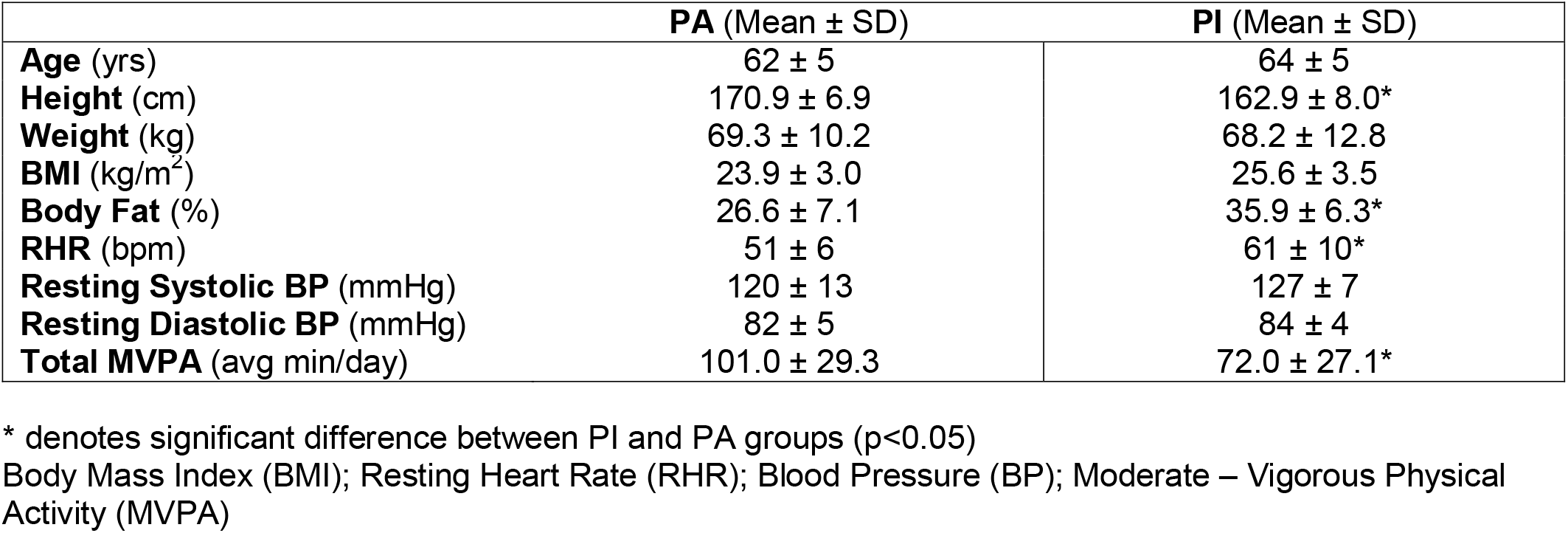
Subject Characteristics. Physically Active (PA; n=12; 6 males, 6 females). Physically Inactive (PI; n=12; 1 male, 11 females).

### Data collection

*Visit 1*. Participants who passed the phone or email pre-screening questions were invited to the Laboratory of Integrated Physiology for assessment. The purpose of the first visit was to ensure participants met inclusion/exclusion criteria, collect descriptive and fitness data, and to familiarize the participants with the exercise equipment and protocols.

Participants filled out a PAR-Q+ and a medical history questionnaire, provided a written list of all medications, and had their seated resting heart rate and blood pressure measured. Height and weight were measured to calculate BMI. This information was reviewed immediately, and if the participant met all inclusion/exclusion criteria then the data collection phase of Visit 1 started.

The eligible participants were classified as being PA or PI,and underwent a Dual Energy X-Ray Absorptiometry (DEXA; Hologic Discovery A DXA, Marlborough, MA, USA) scan to assess body composition. Participants laid supine for 15 minutes, and the lowest HR obtained during this time was used to calculate cardiorespiratory exercise intensity for subsequent laboratory visits using the Karvonen formula [16], with maximum heart rate (MHR) calculated using the formula MHR = 220 - age. Prior to proceeding to the exercise tests, participants were offered a small snack.

### 8-RM Testing

Following a warmup on a motorized treadmill, participants were familiarized with eight stacked weight machine resistance exercises (leg press, chest press, leg curl, lateral pull down, weighted calf raise, triceps extension, leg extension, and seated row).

Participants were instructed on proper form and technique throughout the session, and performed each exercise with minimal weight until they were comfortable with the movement and technique. After this acclimatization, an 8-repetition maximum (8-RM) test was conducted based on the protocols in the American College of Sports Medicine (ACSM) guidelines 10^th^ edition [15]. Each participant performed one set of several repetitions with a light load, after which the resistance was gradually increased until an 8-RM was achieved. The amount of load increase was dependent on each participants’ self-perceived capacity. The Epley formula [17] was used to estimate 1-RM from the 8-RM, and the workload for the RT visit was calculated by taking 70% of estimated 1-RM for each exercise.

### Exercise bout visits

The second and third laboratory visits consisted of CRE or RE, according to the participants’ randomized order. When participants arrived for the exercise bout, study personnel verified that the participant was in a fasted state (at least 8h), that they did not consume alcohol or exercise within the last 24 hours, and that they had not started any new medications.

Participants were fitted with a heart rate monitor and asked to rest in the seated position for 10 minutes, after which time resting heart rate, blood pressure, and a blood sample were collected. All blood samples were collected from a vein in the antecubital space, and collected into sodium heparin-containing tubes for whole blood analyses. The participant then began the exercise bout.

After the 30-minute bout, participants were quickly seated and an immediately post-exercise blood sample was collected. Participants then walked on the treadmill for a cool-down at a slow, self-selected pace for an additional 2 minutes. Following the cool-down, participants rested in the lab for an additional 60 minutes. At 60 minutes post-exercise a third and final blood sample was collected.

### Cardiorespiratory exercise bout

The CRE bout consisted of a 5-minute warm-up of walking at a self-selected pace on the treadmill, followed by an increase in the speed and/or grade of the treadmill such that each participant’s heart rate was within 60-70% of calculated heart rate reserve.

### Resistance exercise bout

The RE bout started with a 5-minute warm-up on a motorized treadmill at a self-selected pace. Following the warm-up, participants completed 3 sets of 8-12 repetitions at 70% of 1-RM with 30-120 seconds of intraset rest and 1-2 minutes of interset rest. All participants completed each exercise in the same order: leg press, chest press, leg curl, lateral pull down, weighted calf raise, triceps extension, leg extension, and seated row. All sets of each exercise were completed before the participant moved onto the next exercise.

### Blood analyses

#### Flow cytometry

Multi-color flow cytometry was used for lymphocyte counts and subsets in whole blood. Flurochrome-conjugated antibodies (Miltenyi Biotech (Bergisch Gladbach, Germany) and a flow cytometer (MACSQuant ® Analyzer 10, Miltenyi Biotech, Gladback, Germany) were used to analyze samples. To determine lymphocyte count, whole blood was labeled with VioBlue-conjugated anti-CD45 mAb, incubated, and treated with RBC lysis buffer. One minute prior to analysis, PI was added and lymphocyte count was determined by forward and side scatter gated on cells within the lymphocyte gate that were CD45+ and negative for PI.

T-cells and T-cell subsets were identified using whole blood labeled with VioBlue-conjugated anti-CD45RA mAb, VioGreen-conjugated anti-CD3 mAb, FITC-conjugated anti-CD4 mAb, PE-conjugated anti-CD196 mAb, PerCP-conjugated anti-CD8 mAb, PE-Vio770-conjugated anti-CD62L mAb, APC-conjugated anti-CD161 mAb, and APC-Vio770-conjugated anti-CD57 mAb. Following a 30-minute incubation, samples were lysed, washed, and analyzed by the flow cytometer. T-cells were identified as those within the lymphocyte gate that expressed the CD3 surface antigen. The CD4+ T-cells (CD3+CD4+) and CD8+ T-cells (CD3+CD8+) were gated within the CD3+ lymphocytes and differentiation status was assessed based on their expression of CD45RA/CD62L and CD57. Th17 cells were identified as those CD4+T-cells that were also CD196+ and CD161+. The total number of T-cell and T-cell subtypes were identified by multiplying the percentage of the cells of interest by the total lymphocyte count.

### Experimental design and statistical analyses

This study was a randomized, complete cross-over design with three time points (pre-exercise, post-exercise, and recovery), two activity levels (PA and PI), and two exercise modalities (cardiorespiratory exercise (CRE), and resistance exercise (RE)). Participants reported to the Laboratory of Integrated Physiology 3 times over the course of the study. All visits were in the morning following an overnight fast. All visits for each participant was scheduled at the same time.

The randomization was stratified by sex, which resulted in equal allocation in the PA group but unequal (5:7) allocation in the PI group. The project biostatistician (KJ) performed the block randomization with random block size, and the project lead (RG) enrolled participants and assigned the intervention. Primary outcome variables were the T-cells, and secondary were the health and fitness measures. Sample size calculations were carried out based on expected differences within inflammatory monocytes (delta = 4.4, SD = 3.7), a primary aim for another aspect of the project. Values related to variability were suggested by previous studies on these same outcomes [18, 19].

Independent sample t-tests were used to determine differences between PA and PI subject characteristics and fitness measurements. To assess baseline differences in immune parameters (differences between PA and PI at pre-exercise), a linear mixed effects model was fit, modeling *condition* (CRE or RE) and *group* (PA and PI) and their interaction.

To determine whether the condition (CRE or RE) influenced the mobilization of T-cells and T-cell subsets as well as to determine whether physical activity status influenced these results, a linear mixed effects model was fit on the change from pre-exercise values with factors *group* (PA and PI), *condition* (CRE and RE), and *time* (pre-exercise, post-exercise, and recovery), along with their interactions. Subject was included as a random intercept term.

Specific comparisons were tested using contrasts. Pre-exercise values served as a linear covariate, and the intercept was tested in order to determine the differences from the pre-exercise time point. Contrasts were used to assess the difference between each groups and types of exercise. Data was transformed using a Box-Cox transformation prior to model fit in order to improve model reliability (i.e. normality and equality of variances). Statistical significance was determined *a priori* at p<0.05. All statistical analyses were conducted with R (version 3.6.2).

## Results

### Participants

The PA group had lower relative body fat (p<0.01) and resting heart rate (RHR; p=0.01), and were taller than the PI group (p=0.03; Table 1). The results from the ActiGraph ® confirmed that PA spent averaged more time per day engaging in moderate to vigorous physical activity (MVPA) compared to PI (p=0.02; Table 2). Furthermore, 1RM’s (Table 2) for PA were greater than PI for leg press (p=0.02), chest press (p=0.01), leg curl (p<0.01), lat pull down (p= 0.01), weighted calf raise (p<0.01), triceps extension (p<0.01), and seated row (p=0.01). Groups did not differ in the calculated target HR at 60% or 70% HRR, leg extension 1RM, nor average exercise HR during the CRE trial. All participants completed both the CRE and RE exercise sessions, but due to difficulties in sampling blood was not collected at the RE recovery time point for two participants in the PI group, and one PI participant in the CRE recovery time point.

**Table 2:**
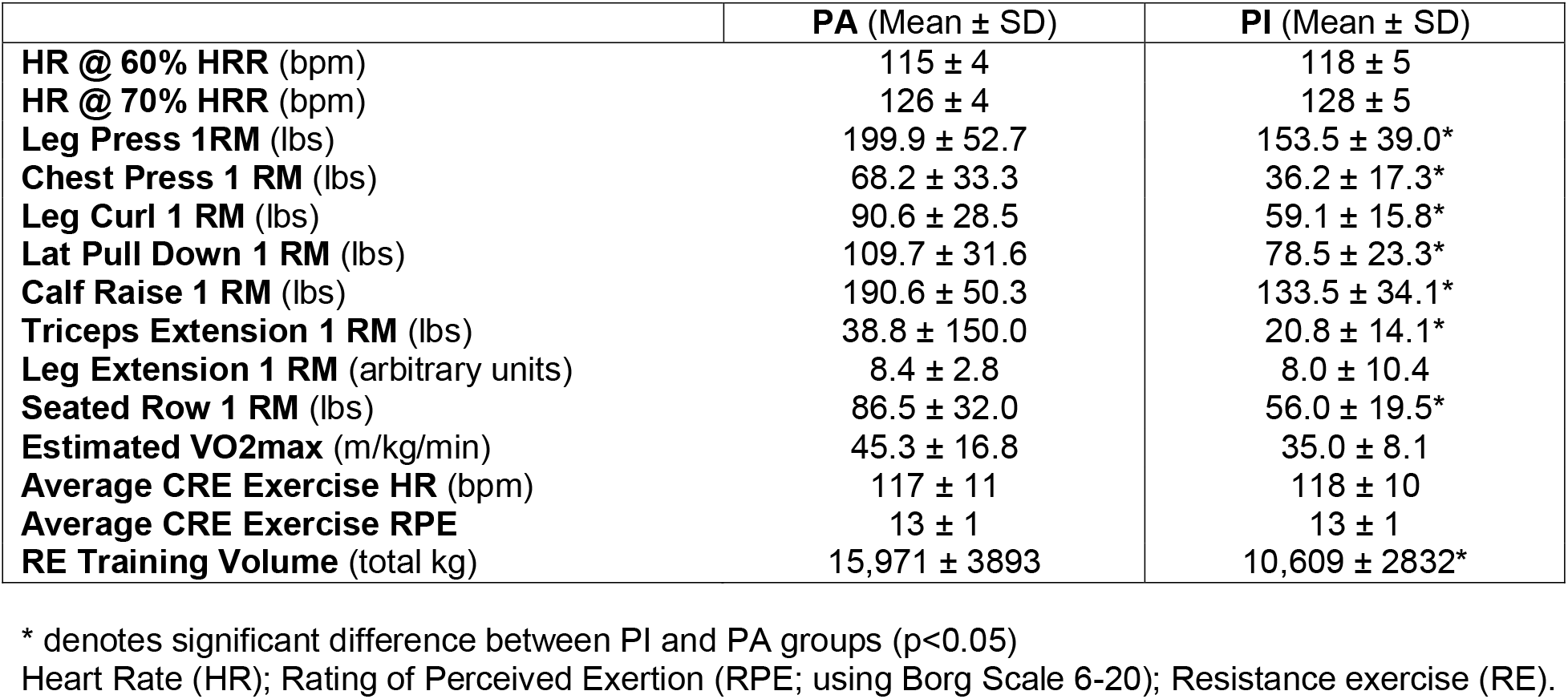
Exercise Data. Physically Active (PA; n=12) and Physically Inactive (PI; n=12) participants completed all study protocols. One Repetition Maximum (RM) values are estimated from 8 RM scores obtained during Visit 1. RE training volume was calculated as sets x reps x weight.

### Pre- to post-exercise T-cell responses

#### Exercise responses that did not differ by group or mode

Irrespective of mode (i.e. exercise conditions CRE and RE not included), exercise mobilized all T-cell subpopulations into the circulation immediately post-exercise in both PA and PI participants (Table 3). When examining response by mode (CRE or RE) of exercise, the only T-cell subpopulation responses that did not differ by group or mode were CD8+ T-cells, EMRA CD8+ T-cells, and CD57+ EMRA CD8+ T-cells. Specifically, the CD8+ T-cells were mobilized independently by CRE and RE exercise in both PA (CRE p=0.006, RE p=0.001) and PI (CRE p=0.046, RE p<0.001) groups (Table 4). EMRA CD8+ T-cells in PA (CRE p<0.001, RE p<0.001) and PI (CRE p=0.004, PI RE p<0.001; Figure 2.A) as well as CD57+ EMRA CD8+ T-cells in PA (CRE p<0.001, PA RE p<0.001) and PI (CRE p<0.001, PI RE p<0.001; Figure 2.B) were also significantly mobilized from pre-exercise to post-exercise.

**Table 3:** Cellular response to exercise training, irrespective of mode of exercise (CRE or RE). Exercise increased the number of all measured cells and subtypes into circulation.

| Cell type | PA (Mean ± SE) | | | PI (Mean ± SE) | | |
| --- | --- | --- | --- | --- | --- | --- |
|  | Pre | Post | p | Pre | Post | p |
| **Lymphocytes** ($\times 10^3$ cells/μL) | 1.3 ± 0.1* | 1.7 ± 0.1 | <0.001 | 1.5 ± 0.1 | 2.0 ± 0.1 | <0.001 |
| **T-cells** ($\times 10^3$ cells/μL)<br>**(CD3+ Lymphocytes)** | 0.7 ± 0.1 | 0.8 ± 0.1 | <0.001 | 0.9 ± 0.1 | 1.1 ± 0.1 | <0.001 |
| **CD4+ T-cells** (cells/μL)<br>**(CD3+ CD4+)** | 486 ± 82 | 523 ± 82 | <0.001 | 692 ± 82 | 798 ± 82 | <0.001 |
| **CD8+ T-cells** (cells/μL)<br>**(CD3+ CD8+)** | 179 ± 26 | 238 ± 26 | <0.001 | 203 ± 26 | 253 ± 26 | <0.001 |
| **Naïve CD4+ T-cells** (cells/μL)<br>**(CD45RA+ CD62L+)** | 128 ± 44 | 131 ± 44 | 0.046 | 265 ± 44 | 289 ± 44 | <0.001 |
| **Naïve CD8+ T-cells** (cells/μL)<br>**(CD45RA+ CD62L+)** | 34 ± 12* | 37 ± 12 | 0.023 | 65 ± 12 | 74 ± 12 | 0.006 |
| **CM CD4+ T-cells** (cells/μL)<br>**(CD45RA- CD62L+)** | 219 ± 33 | 226 ± 33 | 0.002 | 276 ± 33 | 318 ± 33 | <0.001 |
| **CM CD8+ T-cells** (cells/μL)<br>**(CD45RA- CD62L+)** | 26 ± 5 | 30 ± 5 | 0.003 | 33 ± 5 | 38 ± 5 | <0.001 |
| **EM CD4+ T-cells** (cells/μL)<br>**(CD45RA- CD62L-)** | 131 ± 23 | 155 ± 23 | <0.001 | 140 ± 23 | 171 ± 23 | <0.001 |
| **EM CD8+ T-cells** (cells/μL)<br>**(CD45RA- CD62L-)** | 67 ± 12 | 83 ± 12 | <0.001 | 65 ± 12 | 77 ± 12 | <0.001 |
| **EMRA CD4+ T-cells** (cells/μL)<br>**(CD45RA+ CD62L-)** | 7.7 ± 2.8 | 11.3 ± 2.8 | 0.005 | 11.1 ± 2.8 | 19.5 ± 2.8 | 0.006 |
| **EMRA CD8+ T-cells** (cells/μL)<br>**(CD45RA+ CD62L-)** | 52 ± 11 | 89 ± 11 | <0.001 | 40 ± 11 | 63 ± 11 | <0.001 |
| **CD57+ EMRA CD4+ T-cells** (cells/μL)<br>**(CD57+ CD45RA+ CD62L-)** | 1.1 ± 1.5* | 1.7 ± 1.5 | <0.001 | 5.6 ± 1.5 | 9.4 ± 1.5 | <0.001 |
| **CD57+ EMRA CD8+ T-cells** (cells/μL)<br>**(CD57+ CD45RA+ CD62L-)** | 17 ± 5 | 34 ± 5 | 0.007 | 21 ± 5 | 36 ± 5 | 0.007 |
| **Th17 cells** (cells/μL)<br>**(CD4+ CD196+ CD161+)** | 38 ± 10 | 42 ± 10 | 0.004 | 49 ± 10 | 54 ± 10 | 0.002 |

\*Group (PA vs. PI) difference (p<0.05) at pre

**Table 4:** Differences in response by mode of exercise. The effects of an acute bout of cardiorespiratory (CRE) or resistance (RE) exercise on circulating numbers of lymphocytes and lymphocyte subsets in physically active (PA) and physically inactive (PI) participants. Blood samples were taken immediately prior to exercise (Pre), immediately upon exercise cessation (Post) and after 1h of rest following exercise cessation (Recovery) during both CRE and RE trials.

|  |  | PA |  | PI |  |
| --- | --- | --- | --- | --- | --- |
| | | CRE<br>(Mean ± SE) | RE<br>(Mean ± SE) | CRE<br>(Mean ± SE) | RE<br>(Mean ± SE) |
| **Lymphocytes** ($\times 10^3$ cells/μL) | Pre | 1.2 ± 0.2 | 1.3 ± 0.2 | 1.6 ± 0.2 | 1.4 ± 0.2 |
| | Post | 1.7 ± 0.2* | 1.7 ± 0.2* | 2.0 ± 0.2* | 1.9 ± 0.2* |
| | Recovery | 1.1 ± 0.2* | 1.1 ± 0.2* | 1.5 ± 0.2 | 1.4 ± 0.2 |
| **T-cells** ($\times 10^3$ cells/μL)<br>(CD3+ Lymphocytes) | Pre | 0.7 ± 0.1 | 0.7 ± 0.1 | 1.0 ± 0.1 | 0.8 ± 0.1 |
| | Post | 0.9 ± 0.1* | 0.8 ± 0.1* | 1.1 ± 0.1 | 1.1 ± 0.1*^ |
| | Recovery | 0.6 ± 0.1* | 0.7 ± 0.1 | 1.0 ± 0.1 | 0.9 ± 0.1 |
| **CD4+ T-cells** (cells/μL)<br>(CD3+ CD4+) | Pre | 523 ± 87 | 447 ± 87 | 796 ± 87 | 587 ± 87 |
| | Post | 541 ± 87 | 505 ± 87 | 813 ± 87 | 783 ± 87*^ |
| | Recovery | 455 ± 87* | 444 ± 87 | 729 ± 88 | 674 ± 89 |
| **CD8+ T-cells** (cells/μL)<br>(CD3+ CD8+) | Pre | 180 ± 29 | 179 ± 29 | 233 ± 29 | 173 ± 29 |
| | Post | 238 ± 29* | 239 ± 29* | 261 ± 29* | 245 ± 29* |
| | Recovery | 160 ± 29 | 173 ± 29 | 219 ± 29 | 206 ± 30 |
| **Naïve CD4+ T-cells** (cells/μL)<br>(CD45RA+ CD62L+) | Pre | 141 ± 46 | 115 ± 46 | 316 ± 46 | 213 ± 46 |
| | Post | 138 ± 46 | 123 ± 46 | 308 ± 46 | 270 ± 46*^ |
| | Recovery | 117 ± 46* | 112 ± 46 | 268 ± 46 | 225 ± 46 |
| **CM CD4+ T-cells** (cells/μL)<br>(CD45RA- CD62L+) | Pre | 233 ± 35 | 205 ± 35 | 313 ± 35 | 240 ± 35 |
| | Post | 229 ± 35 | 223 ± 35 | 318 ± 35 | 318 ± 35*^ |
| | Recovery | 205 ± 35 | 207 ± 35 | 291 ± 35 | 279 ± 35 |
| **EM CD4+ T-cells** (cells/μL)<br>(CD45RA- CD62L-) | Pre | 141 ± 24 | 120 ± 24 | 154 ± 24 | 125 ± 24 |
| | Post | 160 ± 24* | 150 ± 24* | 166 ± 24 | 176 ± 24*^ |
| | Recovery | 126 ± 24 | 119 ± 24 | 157 ± 24 | 159 ± 24* |
| **EMRA CD4+ T-cells** (cells/μL)<br>(CD45RA+ CD62L-) | Pre | 8 ± 3 | 7 ± 3 | 13 ± 3 | 9 ± 3 |
| | Post | 13 ± 3* | 9 ± 3 | 21 ± 3* | 18 ± 3* |
| | Recovery | 7 ± 3 | 6 ± 3* | 13 ± 3 | 11 ± 3 |
| **CD57+ EMRA CD4+ T-cells**<br>(cells/μL)<br>(CD57+ CD45RA+ CD62L-) | Pre # | 1 ± 2 | 1 ± 2 | 7 ± 2 | 5 ± 2 |
| | Post | 2 ± 2* | 2 ± 2 | 9 ± 2 | 10 ± 2*^♦ |
| | Recovery | 1 ± 2 | 1 ± 2 | 6 ± 2 | 6 ± 2 |

\* within-group difference compared to pre-exercise ( $p < 0.05$ )
^ within-group differences from the same time point in CRE compared to RE ( $p < 0.05$ )

**Figure 2:**
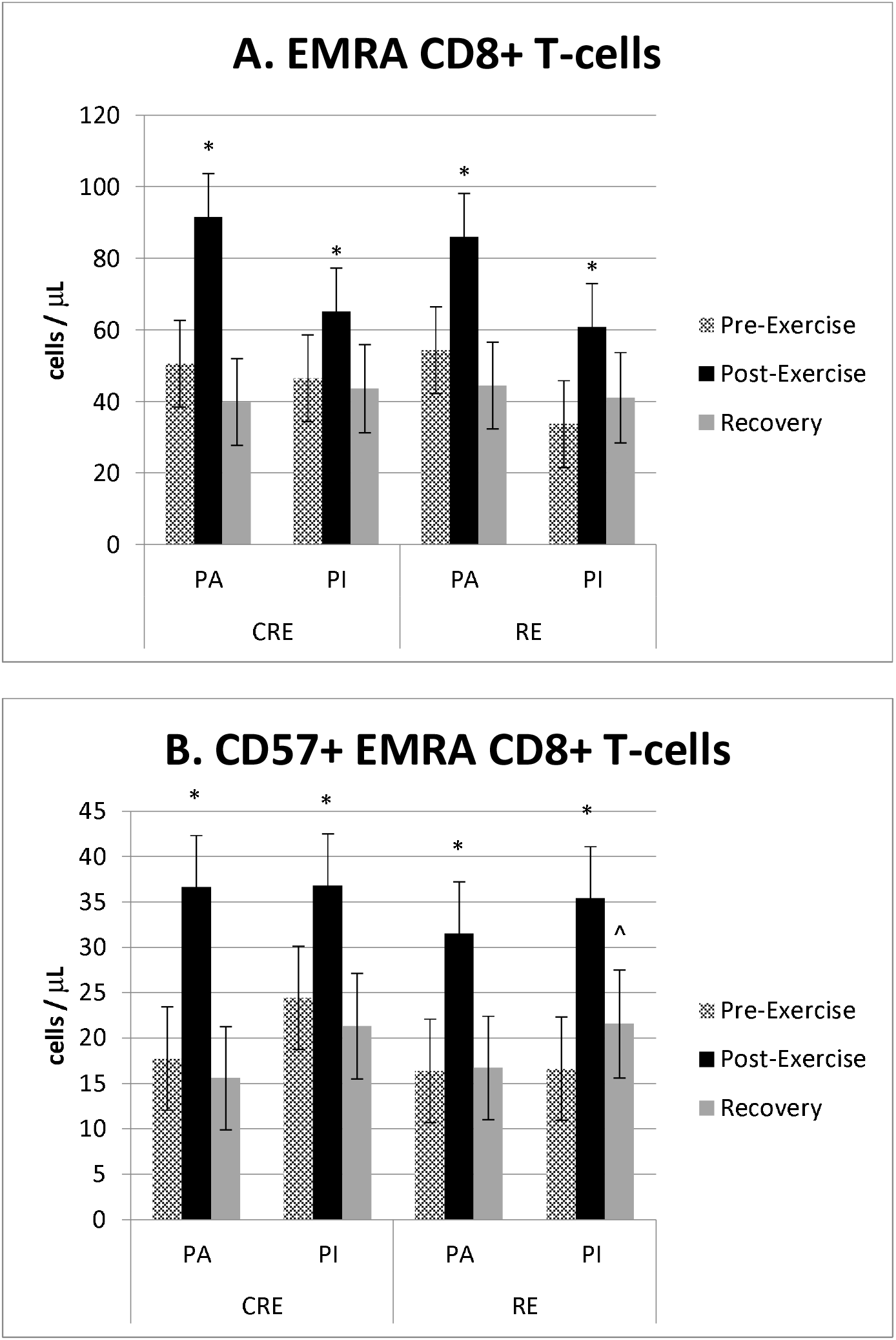
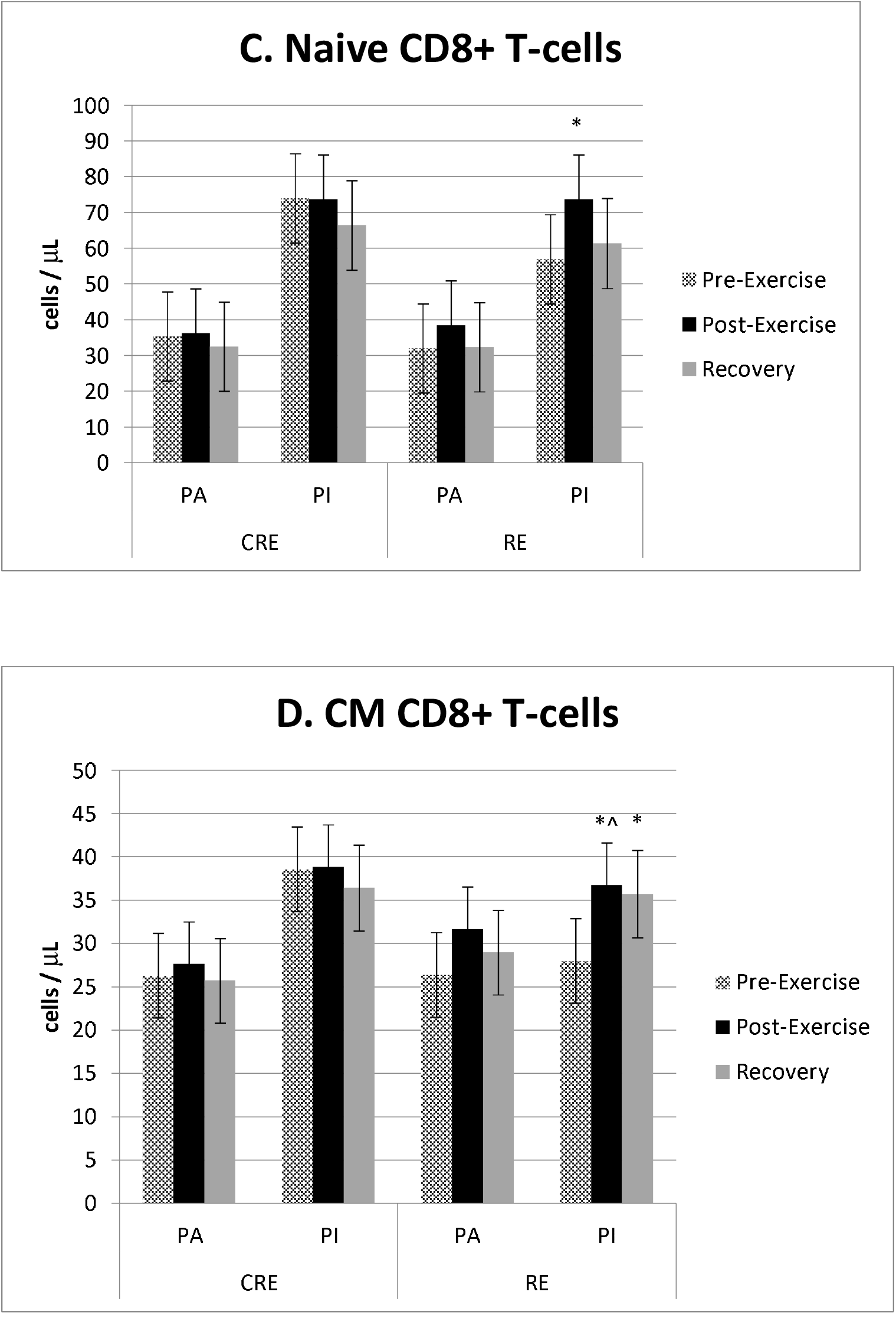
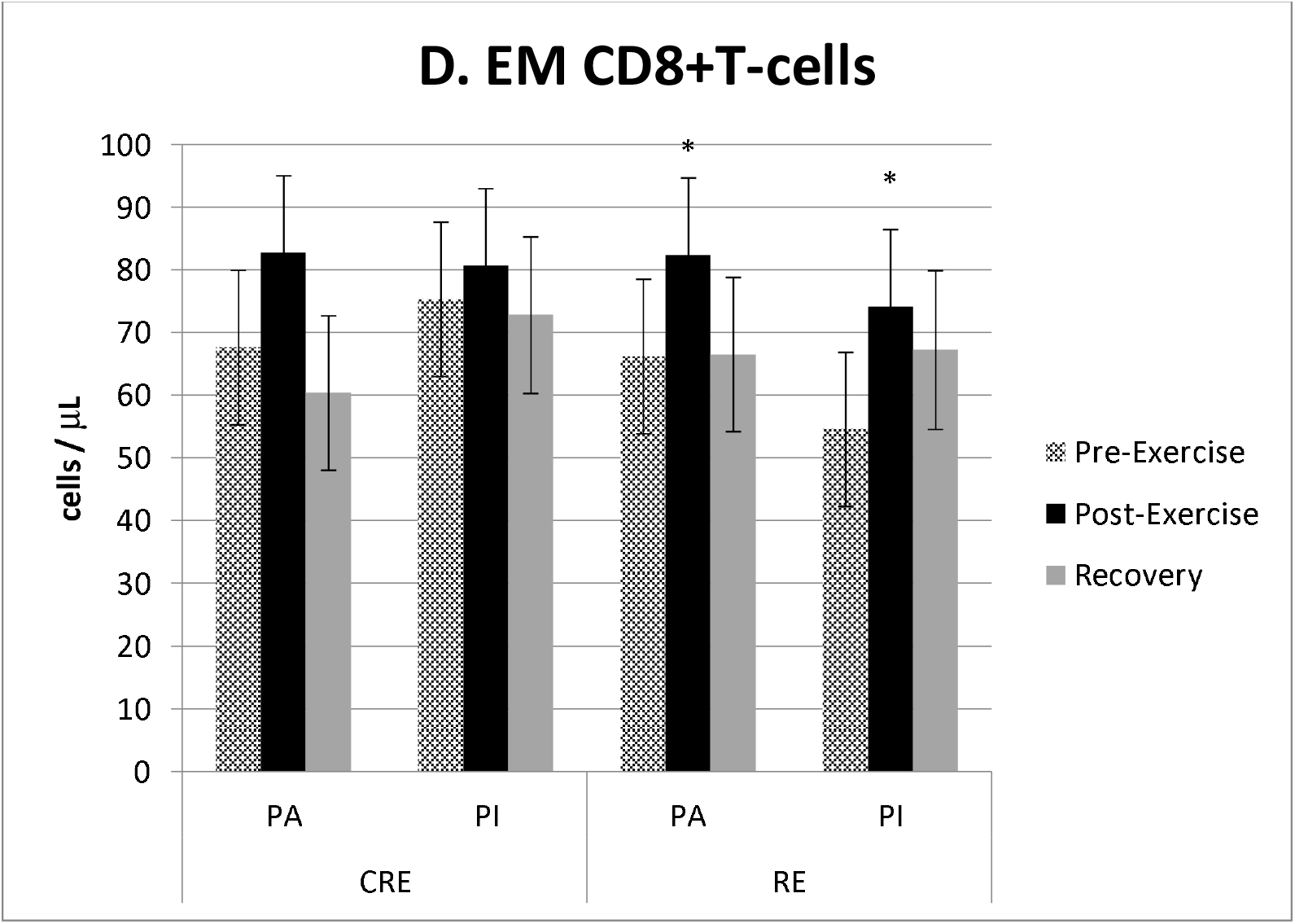
CD8+T-cell subsets at pre-exercise, post-exercise, and recovery in response to cardiorespiratory (CRE) and resistance (RE) exercise in physically active (PA) and physically inactive (PI) groups. * difference between pre- to post-exercise (p<0.05) ^ within-group differences from the same time point in CRE (p <0.05)

### More T-cell subpopulations mobilized pre- to post-exercise in RE than CRE in PI group

Within PI, all circulating T-cells and T-cell subpopulations increased in number from pre-exercise to post-exercise with RE, but only five of the measured populations and subpopulations increased in both RE and CRE. The cells that increased in circulation immediately post-exercise in response to RE but not CRE were T-cells (p<0.001, Table 4), CD4+ T-cells (p<0.001, Table 4), naïve CD4+ T-cells (p<0.001, Table 4), naïve CD8+ T-cells (p=0.002, Figure 2.C), CM CD4+ T-cells (p<0.001, Table 4), CM CD8+ T-cells (p=0.001, Figure 2.D), EM CD4+ T-cells (p<0.001, Table 4), EM CD8+ T-cells (p=0.003, Figure 2.E), CD57+ EMRA CD4+ T-cells (p<0.001, Table 4), and Th17 cells (p=0.004, Figure 3). Conversely, both RE and CRE significantly mobilized total lymphocytes (RE p<0.001, CRE p<0.001, Table 4), EMRA CD4+ T-cells (RE p<0.001, CRE p<0.001, Table 4), and the aforementioned CD8+ T-cells, EMRA CD8+ T-cells, and CD57+ EMRA CD8+ T-cells.

**Figure 3:**
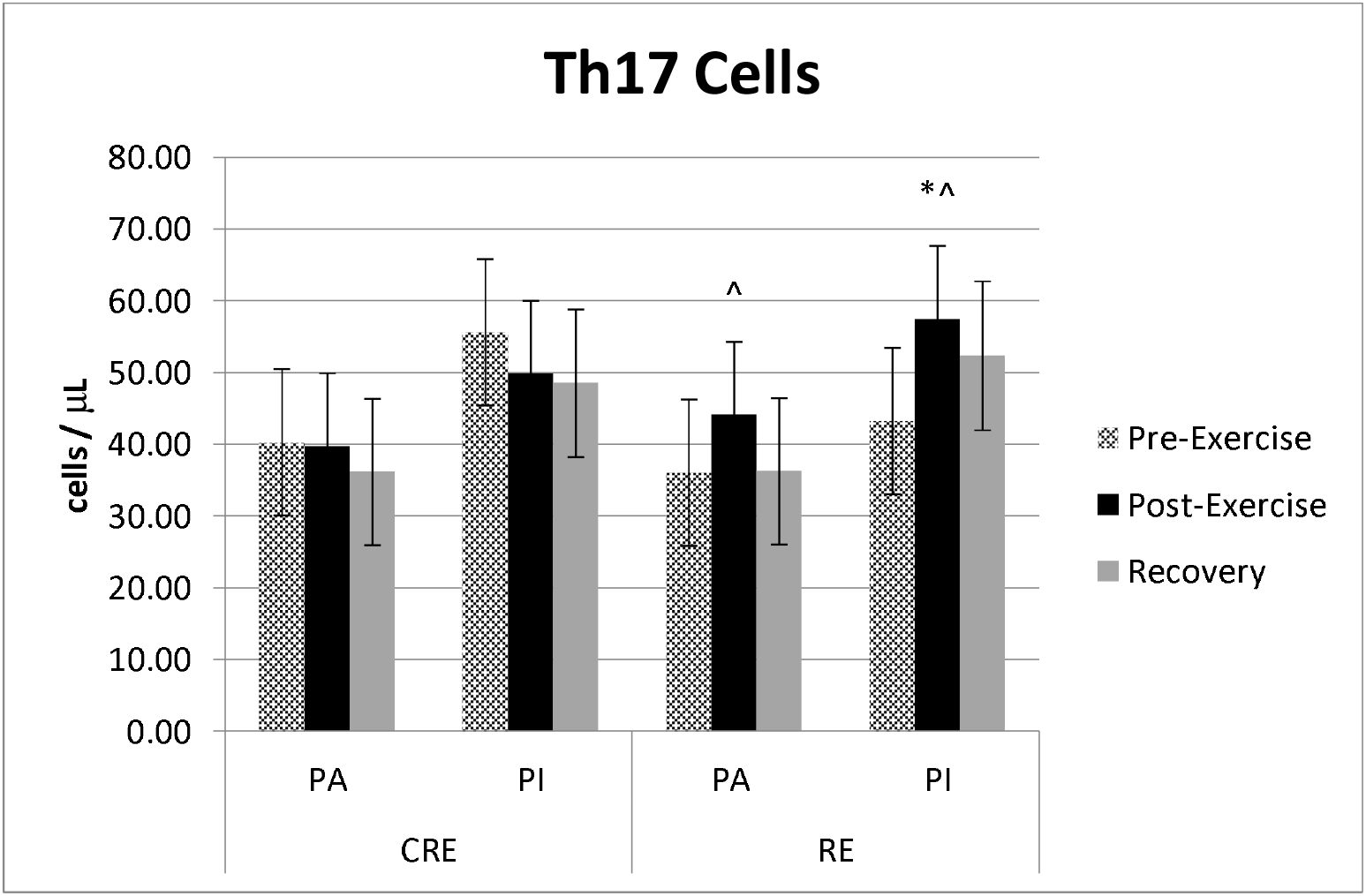
Th17 cells response. Th17 Cells at pre-exercise, post-exercise, and recovery in response to cardiorespiratory (CRE) and resistance (RE) exercise in physically active (PA) and physically inactive (PI) groups * Difference compared to pre values (p< 0.05) ^ Within-group differences from post-exercise CRE (p <0.05)

Within PA, unlike PI there were no cell populations that were mobilized only by RE. CRE selectively mobilized EMRA CD4+ T-cells (p<0.001, Table 4) and CD57+ EMRA CD4+ T-cells (p=0.020, Table 4). Also unlike PI, both RE and CRE mobilized T-cells (RE p=0.001, CRE p=0.006, Table 4).

### Exercise recovery response in T-cells

#### Some T-cell subtypes remain elevated at recovery time point in the PI group

Within the PI group only, the EM CD4+ T-cells (p=0.006, Table 4) and CM CD8+ T-cells (p=0.025, Figure 2.D) remained elevated above pre-exercise values at the RE recovery time point. Recovery was not higher than immediately post-exercise. All T-cells and T-cell subtypes returned to pre-exercise counts at the recovery time point within the PI group with CRE exercise.

#### Some T-cells decrease at the recovery time point in the PA group

Conversely, in the PA participants, RE caused a decrease from pre-exercise during the recovery time point for total lymphocytes (p=0.002, Table 4) and EMRA CD4+ T-cells (p=0.035, Table 4), while CRE decreased recovery total lymphocytes (p<0.001, Table 4), T-cells (p=0.037, Table 4), CD4+ T-cells (p=0.029, Table 4), and naïve CD4+ T-cells (p=0.005, Table 4) below pre-exercise.

## Discussion

This is the first study that used a crossover design to examine the T-cell response in older physically active and inactive adults who completed separate bouts of acute moderate-intensity CRE and RE. We report that regardless of physical activity status, moderate-intensity CRE and RE both facilitate a significant mobilization of the highly-differentiated EMRA CD8+ T-cells and CD57+ EMRA CD8+ T-cells into the circulation. Further, the Th17 cells response is similar between PA and PI groups, but differs by mode of exercise (CRE compared to RE). Moreover, within the PI group RE mobilized more T-cell subpopulations than CRE. We also confirmed that older adults who are physically active have lower numbers of circulating senescent CD57+ EMRA CD4+ T-cells compared to age-matched physically inactive adults.

Conventional hypotheses on declines in immunity link aging and immunosenescence, a canopy term used to describe the age-related decline in immune competency. For example, one characteristic of age-related immune decline is an accumulation of highly-differentiated T-cells and a concomitant reduction in the naïve T-cell pool [6]. However, physical activity has recently emerged as a potentially more potent mediator of the relationship between aging and immune decline [2]. This newer hypothesis of physical activity as the mediating factor of age-related immune decline is supported by several cross-sectional and longitudinal studies, which demonstrate a marked difference in immune characteristics of those that engage in regular physical activity compared to those who do not. Further, as physically inactive adults engage in regular exercise training and improve their physical fitness, several immune parameters are altered in a way that more closely mirrors those of a physically active adult [6, 7, 10, 11].

Specifically, older adults who participated in a combined resistance and cardiorespiratory exercise training program increased their CD28+CD8+ T-cells, yet the participants in the control group had no change in this cell subset [11].

Results from the present study partially support the earlier findings of the effect of physical activity on age-associated immune decline. Specifically, highly differentiated CD4+ T-cells (CD57+ EMRA CD4+ T-cells) were higher in PI compared to PA at rest (pre-exercise). This study also confirmed that regardless of exercise mode of physical activity level of the participants, exercise mobilizes highly-differentiated CD8+ T-cells. These highly-differentiated CD8+ T-cells are known to have heightened effector functions compared to their lower-differentiated and CD4+ T-cell counterparts [9, 20, 21]. Contributing to this knowledge of exercise-induced mobilization, the present study adds that the significant mobilization of highly-differentiated CD8+ T-cells can occur with moderate intensity CRE and RE in a population of older adults who are physically active or physically inactive. Therefore, the exercise-induced mobilization of highly-differentiated CD8+ T-cells is non-discriminate. However, the intensity of exercise in this study was the same in both exercise modes. Future studies should explore the contribute of intensity of exercise in both CRE and RE to the mobilization of highly-differentiated CD8+ T-cells.

There are several potential benefits associated with the mobilization of highly-differentiated CD8+ T-cells, along with other leukocytes with increased effector functions, inflammatory properties, and cell adhesion markers (specifically, NK cells, gamma delta T-cells, and non-classical monocytes). For example, some researchers speculate that the acute exercise-induced mobilization and rapid egress of these effector cells can heighten the body’s ability to fight infection [7]. It is hypothesized that exercise serves as the initial stimulus to recruit these immune cells with specific properties into the circulation, and then rapidly redistribute them to mucosal surfaces where they will facilitate heightened immune surveillance—thereby enhancing the body’s ability to launch an immune response against an invading antigen [7]. This is in contrast to the widely discussed “open-window theory”, which postulates that the egress of leukocytes within the hours and days following acute exercise serves as a period of immunosuppression during which the host is at an increased risk for opportunistic infection [22]. The evidence to support the open window theory is waning; conversely, the evidence supporting a post-acute exercise elevated state of immunity is mounting [7]. Thus, regular exercise may serve to heighten immunity by allowing an individual to have repeated periods of heightened immunity more frequently than an individual who does not exercise.

Regular exercise and therefore frequent mobilization and subsequent egress of highly-differentiated CD8+ T-cells may provide additional benefits to the overall properties of the immune system. A hallmark of immunosenescence is an accumulation of highly-differentiated T-cells accompanied by a reduction in the naïve T-cell pool, which hypothetically places an individual at a heightened risk of novel infections due to a reduction in the ability of the immune system to respond to a novel antigen [23]. It has been proposed that the exercise-induced recruitment of highly-differentiated T-cells into the circulation both facilitates an increased opportunity for apoptosis (i.e. the removal of excess memory T-cell clones) and simultaneously stimulates thymic output, resulting in the production of naïve T-cells [23]. Indeed, the results from several studies support that apoptosis following acute exercise is increased [24], and also that although thymic involution begins earlier in life (i.e. around puberty), the thymus continues to produce new T-cells well into advanced aging [25] and exercise may serve as a means of stimulating an increase in thymic output [23]. In summary, the immunological disturbances to the T-cell pool induced by regular bouts of exercise may serve to improve immunity in older adults via several physiological pathways.

In contrast to the group differences in highly differentiated CD4+ T-cells, there were not consistent differences between the physical activity groups for naïve T-cells within either the CD4+ or the CD8+ subpopulations at rest (pre-exercise). This is in agreement with a published study comparing Masters Athletes and age-matched controls [26]. In the present study, the only exercise-induced increase in naïve T-cells was in the PI group in response to RE. It is possible that a lower or higher intensity of exercise, or longer or shorter mode of exercise may be required to elicit a naïve T-cell response in more subpopulations or in physically active adults.

This is the first study to report a response of Th17 cells to moderate intensity CRE and RE in a cohort of older adults. There was a differential response of Th17 cells; circulating numbers of Th17 cells were increased in response to RE, yet decreased in response to CRE. Th17 cells are a subset of T-helper (CD4+) cells that produce large amounts of IL-17, a pro-inflammatory cytokine that is implicated in allergic reactions and autoimmunity [27].

Concentrations of Th17 cells are also reported to increase with age [28], and may be mitigated with regular physical activity [29]. However, very little is known about the response of Th17 cells to acute exercise, especially in humans. Limited data supports that Th17 cells increase in response to ultra-endurance or exhaustive exercise events [30, 31], and that this response may be stimulated by changes in serum cytokine levels [30]. However, it must be noted that methodological differences in the ways of identifying Th17 cells make it difficult to compare results. Future research should aim to determine the mechanisms of mobilization of these cells and potential reasons why the response may be different among different training modalities.

The findings reported in this study are particularly intriguing, considering most other immune cell subsets appear to respond similarly between CRE and RE.

Although the immune response to CRE and RE in the present study appear to be similar, some marked differences were found within the PI group in particular. All T-cells and T-cell subtypes measured were significantly elevated post-exercise following RE but not CRE. This is likely because RE served as a novel stimulus for those in the PI group; even though RE was performed at a moderate intensity, it still served as a physiological stimulus that the body was unaccustomed to responding to, which may have resulted in the observed more robust reaction. Indeed, research comparing trained vs. untrained individuals supports that the absolute number of cells mobilized into the circulation following an acute bout of exercise is lower in those who are trained compared to untrained individuals [13]. As the CRE exercise consisted of treadmill walking, it is reasonable that the immune response of the PI group was closer to that of their PA counterparts because even though regular, structured physical activity was not a lifestyle habit employed by those in the PI group, walking did not serve as a novel stimulus to these individuals.

This novel exercise premise is reinforced by the data from the recovery time point, whereby a few T-cell subsets (EM CD4+ T-cells, and CM CD8+ T-cells) remained significantly elevated above pre-exercise levels following RE in the PI group only. This is in contrast to the PA group, where all T-cell subtypes were either back at pre-exercise or below pre-exercise values at the recovery time point. Such group differences suggest a reduction in the efficiency of the immune response between PA and PI. These physical activity group differences may be attributed to a number of differences observed in physically active vs. inactive people, including differences in β2-adrenergic receptor sensitivity, glucocorticoid receptor sensitivity, or cell adhesion molecules [13].

More research is necessary in order to delineate the precise frequency, intensity, modality, and duration of activity necessary to facilitate these improvements [7]. This is illustrated in a study where older adults (65-85 years) were categorized as never trained (NT), leading a moderate training lifestyle (MT), and leading an intense training lifestyle (IT).

Differences between the number of senescent T-cells (using cell surface markers CD45RA and CCR7) were found only between the NT and IT groups [32]. Furthermore, there were no differences in CD28^-^ T-cells between any of the groups [32]. Together, these results support that there may be both a dose-type response in which regular physical activity exerts anti-aging immunological changes and also that the benefits of regular exercise on immunity may not be readily apparent among all ways of characterizing markers of immunosenescence. Specifically, within the same group of participants, significant differences were found in the proportions of highly differentiated t T-cells between physically active and inactive individuals when using cell surface markers CD45RA and CCR7. But, within the same participants, no significant differences were found in the proportions of T-cells when using CD28 as a senescence marker.

The are several items to note in regards to the methodologies for this study. For example, the inclusion / exclusion criteria for the study were quite rigorous, such that the PI group consisted of a “healthy” group of older individuals, without many of the comorbidities that often accompany aging (i.e. diabetes, obesity, cardiovascular conditions, and those who were taking beta blockers for high blood pressure or statins for high cholesterol). This was done in an attempt to focus on the effects of regular physical activity on immune parameters in the absence of obvious confounders and serves as a major strength of the study. Furthermore, we used the ACSM guidelines for physical activity and participants’ self-reported activity in order to stratify between our physically active and inactive groups. This means that the gap between physically active and inactive was not tremendous (i.e. the active group did not consist of competitive athletes, while the inactive group was otherwise quite healthy and while they did not engage in regular, structured exercise, they did lead ambulatory lifestyles), and thus more research is needed in order to determine whether there are further differences in immune characteristics and responses in those who engage in higher volumes and intensities of physical activity compared to those who are even more sedentary. Additionally, latent CMV infection was not controlled for in the present study and is known to affect the composition of the T-cell pool and T-cell responses to acute exercise. However, this was addressed in part statistically by using each participant’s pre-exercise value as a covariate in the model. Lastly, the PA group consisted of an equal number of male and female participants, whereas PI consisted primarily of female participants, which may have influenced group differences [33].

In conclusion, this is the first study to compare moderate intensity CRE and RE in a cohort of physically active and inactive older adults of otherwise similar health status. Our findings indicate that overall CRE and RE have similar effects on T-cell subset mobilization, including a significant mobilization of highly-differentiated CD8+ T-cells, but there were some differences—notably, the response of Th17 cells. As the majority of existing literature on exercise and the immune response is derived from cardiorespiratory exercise studies, these findings provide important insight into the effects of resistance training on the immune response to exercise.

## Supporting information

CONSORT checklist

## Data Availability

Data available from the corresponding author by request

